# Protection Level and Reusability of a Modified Full-Face Snorkel Mask as Alternative Personal Protective Equipment for Healthcare Workers During the COVID-19 Pandemic

**DOI:** 10.1101/2020.08.16.20176081

**Authors:** Jean Schmitt, Lewis S. Jones, Elise A. Aeby, Christian Gloor, Berthold Moser, Jing Wang

## Abstract

The worldwide outbreak of the COVID-19 drastically increased pressure on medical resources and highlighted the need for rapidly available, large-scale and low-cost personal protective equipment (PPE). In this work, an alternative full-face mask is adapted from a modified snorkel mask to be used as PPE with two medical grade filters and a 3D-printed adapter. As the mask covers the eyes, mouth and nose, it acts as a full-face shield, providing additional protection to healthcare workers. The filtration efficiency of different medical filters is measured for particles below 300 nm to cover the size of the SARS-CoV-2 and small virus-laden droplets. The filtration performance of the adapted full-face mask is characterized using NaCl particles below 500 nm and different fitting scenarios. The mask is compared to a commercial respirator and characterized according to the EN 149 standard, demonstrating that the protection fulfills the requirements for the FFP2 level (filtering face-piece 2, stopping at least 94% of airborne particles). The device shows a good resistance to several cycles of decontamination (autoclaving and ethanol immersion), is easy to be produced locally at low cost and helps addressing the shortage in FFP2 masks and face shields by providing adequate protection to healthcare workers against particles below 500 nm.

## INTRODUCTION

The outbreak of COVID-19 in 2019 and the subsequent global spread through 2020 caused an overload of the healthcare system in many affected countries. Hospitals and medical facilities faced a shortage of personal protective equipment (PPE), highlighting the need for rapidly available and large-scale emergency solutions 1. With developing countries facing similar issues, low-cost and local availability are also important characteristics of the required PPE. Therefore, several research groups have worked on adapting and testing existing devices likely to be used as PPE, *e.g*., the use of charged nanofibers to filter SARS-CoV-2 [2] and the successful adaptation of an elastomeric respirator as an alternative to the N95 respirators [3]. Several projects are focusing on the filtration efficiency of a snorkel full-face mask modified with an open-source 3D-printed adapter. A study from [4] covered the filtration efficiency in the range 0.3–10 µm together with the measurement of carbon dioxide accumulation and sound propagation through the mask. Another publication [5] performed a successful fit test with a similar mask, fulfilling the OSHA N95 standard.

The SARS-CoV-2 has been measured between 60 nm and 140 nm [6] and aerosols containing SARS-CoV-2 are mostly found in the 250–500 nm size range [7]. Particles below 10 µm generated by a coughing patient are likely to travel over several meters before settling on the floor [8]. A significant number of particles generated during expiratory activities are smaller than 800 nm [9]. The transmission of COVID-19 through virus-laden droplets is actively investigated [10, 11]. The most penetrating particle size (MPPS) corresponds to the size where the filters have their minimum efficiency and results from the combined effects of the filtration mechanisms (interception, inertial impaction, diffusion and electrostatic attraction) at different particle sizes. The MPPS is usually in the range from 100 to 300 nm for mechanical filters and between 30 and 70 nm for electrostatic filters [12]. Therefore, measuring the penetration of particles with a diameter below 500 nm is critical as they are in the range where filters have their lowest efficiency, and may be highly contagious. This is because these particles exhibit high mobility and a long suspension time. The treatment of infected patients often involves aerosol-generating medical procedures, such as intubation, bronchoscopy and mechanical ventilation, and increases the generation of potentially contaminated droplets [13]. The adapted full-face mask tested in this work prevents contaminated droplets from reaching the eyes, mouth and nose of medical staff performing such procedures. The adaptation of full-face snorkel masks with locally available filters and easy-to-produce adapters can constitute an alternative source of PPE to overcome the shortage in FFP2 masks and face shields.

The aim of the present work is to provide new data on modified snorkel masks, similar to those described by [4] and [5], with a novel adapter designed to hold two medical filters. The focus is set on the size-resolved particle penetration below 500 nm as well as the influence of decontamination processes on the protection efficiency. In the first section, the filtration efficiency of several types of filters is measured and the quality factor is calculated based on filter pressure drop and filtration efficiency at the MPPS for particles between 12 nm and 300 nm. In the second part, the size-dependent particle penetration, from 15 nm to 500 nm, in the full-face mask with the adapter and filters is measured whilst placed on a dummy head. Different fitting scenarios are considered and a commercial full-face respirator is tested as a reference. Finally, the impact of two decontamination methods (autoclaving and ethanol immersion) on the protection efficiency is measured.

## RESULTS AND DISCUSSION

### Description of the full-face mask and adapter

Figure 1 presents the snorkel mask and adapter. The mask is divided into two chambers, with the upper one enclosing the eyes (marked 1) and the lower one enclosing the mouth and nose (marked 2). The chambers are separated by two unidirectional valves (marked 3), which open during inhalation (Figure 1b) and close during exhalation (Figure 1c). The chin port (marked 4) also contains a unidirectional valve. Due to the potential expulsion of contaminated droplets with exhalation, the chin port is permanently sealed with a disk of PMMA. Sealing slightly reduces particle penetration during inhalation and does not appear to cause any increase in humidity or temperature. Further details can be found in the supporting information (Figure S1).

**Figure 1.**
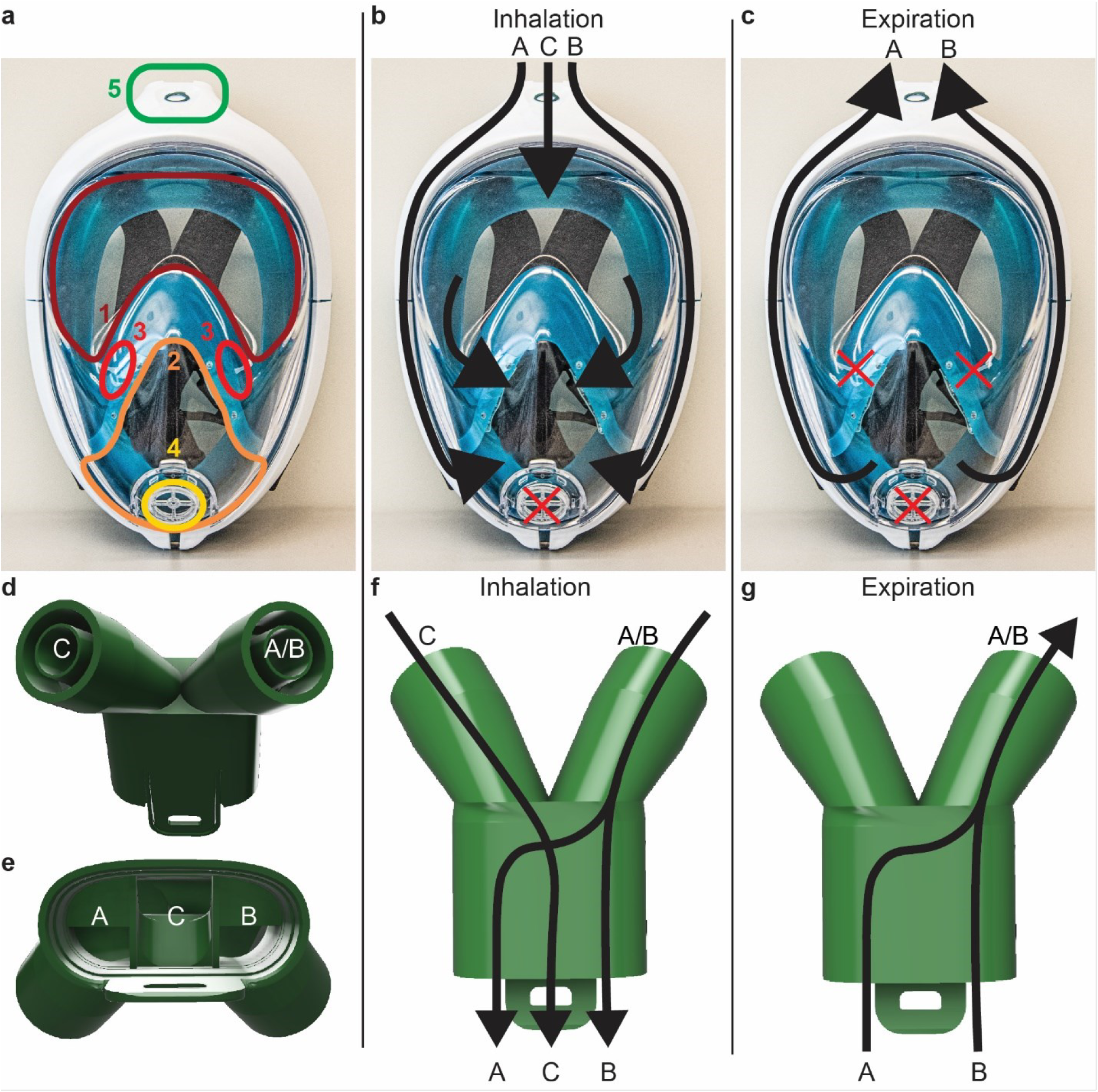
The snorkel mask and 3D-printed adapter. (a) shows the subdivisions of the mask: the upper chamber (marked 1) encloses the eyes and lower chamber (marked 2) the mouth and nose. They are separated by a piece of silicone holding two unidirectional valves (marked 3). The lower chamber contains the unidirectional chin valve (marked 4). The port for the adapter is located on the top (marked 5). (b) shows the airways during inhalation, as the air enters from all three inlets on top, marked A, B and C, (c) shows the airways during exhalation (with a sealed chin valve) only involving the side channels, marked A and B. (d) and (e) show the 3D-printed adapter, the two medical filters are connected to the ports marked A/B and C in (d). The airflows through the adapter are shown in (f) for the inhalation and in (g) for the exhalation. The channels A and B merge into one channel (marked A/B) inside the adapter.

The 3D printed adapter, shown in Figure 1d-g, is adapted from an existing open-source design to enable the fitting of two filters. The filters and mask are connected by friction fit. The adapter is designed such that the attached filters are angled towards the back of the wearer’s head; this provides protection against frontally projected airborne particles [14] and is ergonomic. The adapter directs air inflow from the filters via a central channel (Fig. 1b, marked C) into the upper chamber of the mask, or via two lateral channels (Fig. 1b, marked A & B) into the lower chamber. This allows inhaled air to enter the mask through both filters, whilst exhaled air is expulsed only via the lateral channels of the mask and one filter. The one-way valves of the mask enable this flow separation, which reduces the breathing resistance of the assembly.

### Tests under real operating conditions

The full-face mask was tested for several days in a medical facility during the peak of the SARS-CoV-2 outbreak in Switzerland. It was worn for up to 16 h a day. An adapter with one filter (all three channels merged into one and led to one filter) was compared to the adapter used in this work. The breathing was reported to be easier with the two-filter adapter, leading to improved comfort over long periods of time. A seal check has been performed and the straps have been adapted before using the mask. The volunteers described a feeling of safety and protection brought by the full coverage of the face. The mask is also easy to handle with gloves, lowering the risk of contamination during the installation and removal of the full-face mask. However, the configuration of the face mask does not allow wearing glasses together with the mask.

### Filter selection

Different filters used for mechanical ventilation in intensive care units and operation rooms are compared to each other and to FFP2 and FFP3 (respectively filtering face-piece 2 and 3, corresponding to a filtration efficiency of at least 94% and 99% according to EN 149 standard) filtration materials taken from commercial respirators. The objective of the comparison is to select the filters with the highest efficiency for subsequent measurements: as the focus of this study is to characterize the full-face mask, the filter itself should have a limited influence on the measurements of the particle penetration into the mask. The experimental setup is presented in Figure 2a and the particle penetration for each measured size at a constant face velocity of 8.5 cm/s is given in Figure 2b.

**Figure 2.**
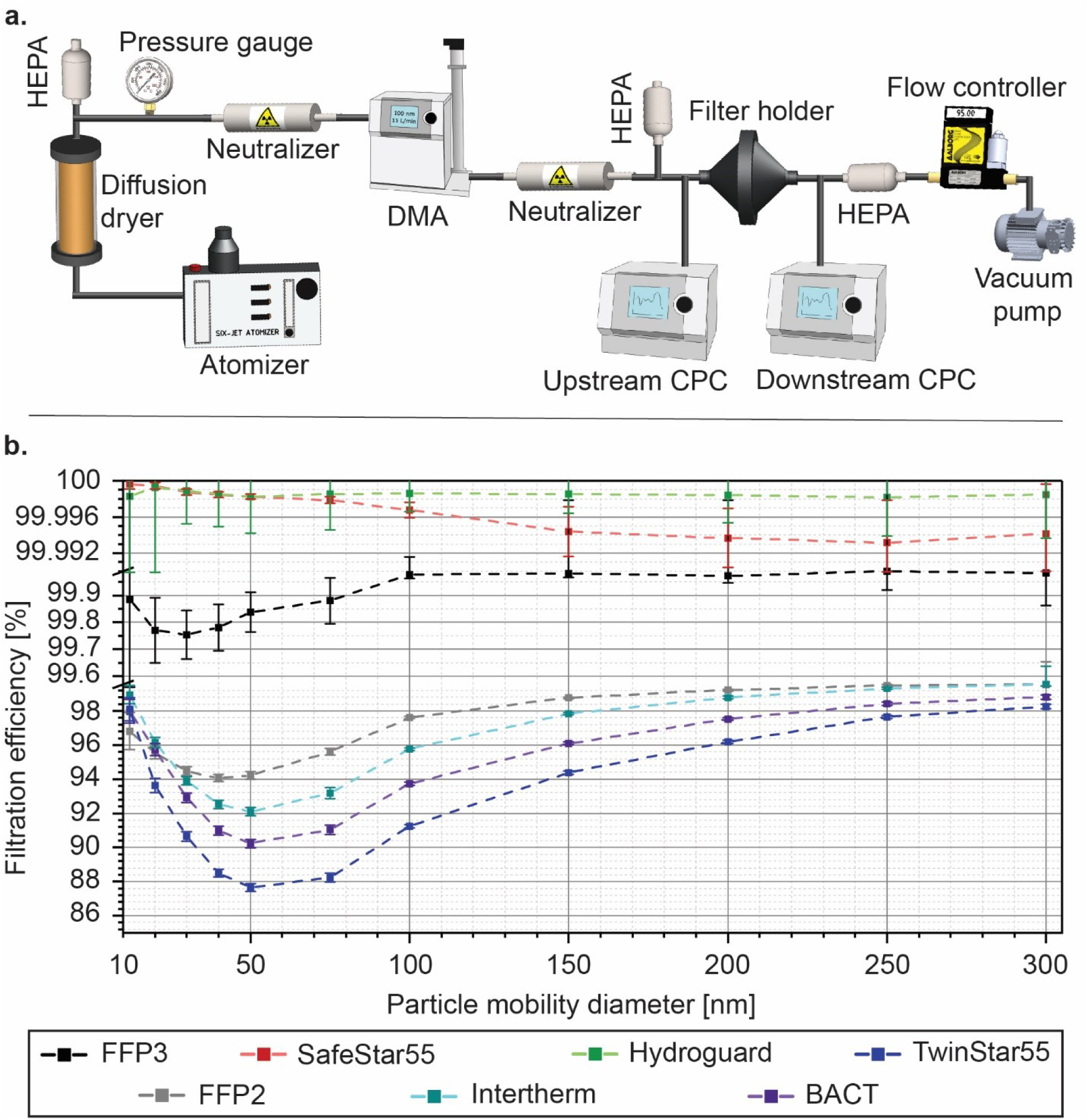
Measurement of the filtration efficiency of five medical grade filters and comparison with filtration media from FFP2 and FFP3 face masks. (a) presents the experimental setup to measure the penetration of NaCl particles with different mobility diameters. The measurement is performed with a differential mobility analyzer (DMA) and two condensation particle counters (CPC). (b) shows the filtration efficiency as a function of the particle mobility diameter at a constant face velocity of 8.5 cm/s. The error bars refer to the standard deviation calculated from the 300 measurements points. The sampling time is 5 min.

The SafeStar55 and Hydroguard, both labelled as mechanical HEPA (high efficiency particulate air) filters, have the best filtration efficiency, over 99.99%. Their most penetrating particle size (MPPS), which corresponds to the minimum filtration efficiency, is around 250 nm. The other filters, based on electrostatic filtration, have a smaller MPPS between 30 nm and 70 nm and the shape of the curve describing the filtration efficiency as a function of the particle size is similar to the FFP2 and FFP3 filter materials. Their efficiency is over 98% at 300 nm. The filtration efficiency and MPPS measured for the FFP2 and FFP3 masks are similar to those measured by [15].

The measurements of the pressure drop generated by the full-face mask equipped with the adapter and the filters are presented in Figure 3 together with the calculation of the quality factor.

**Figure 3.**
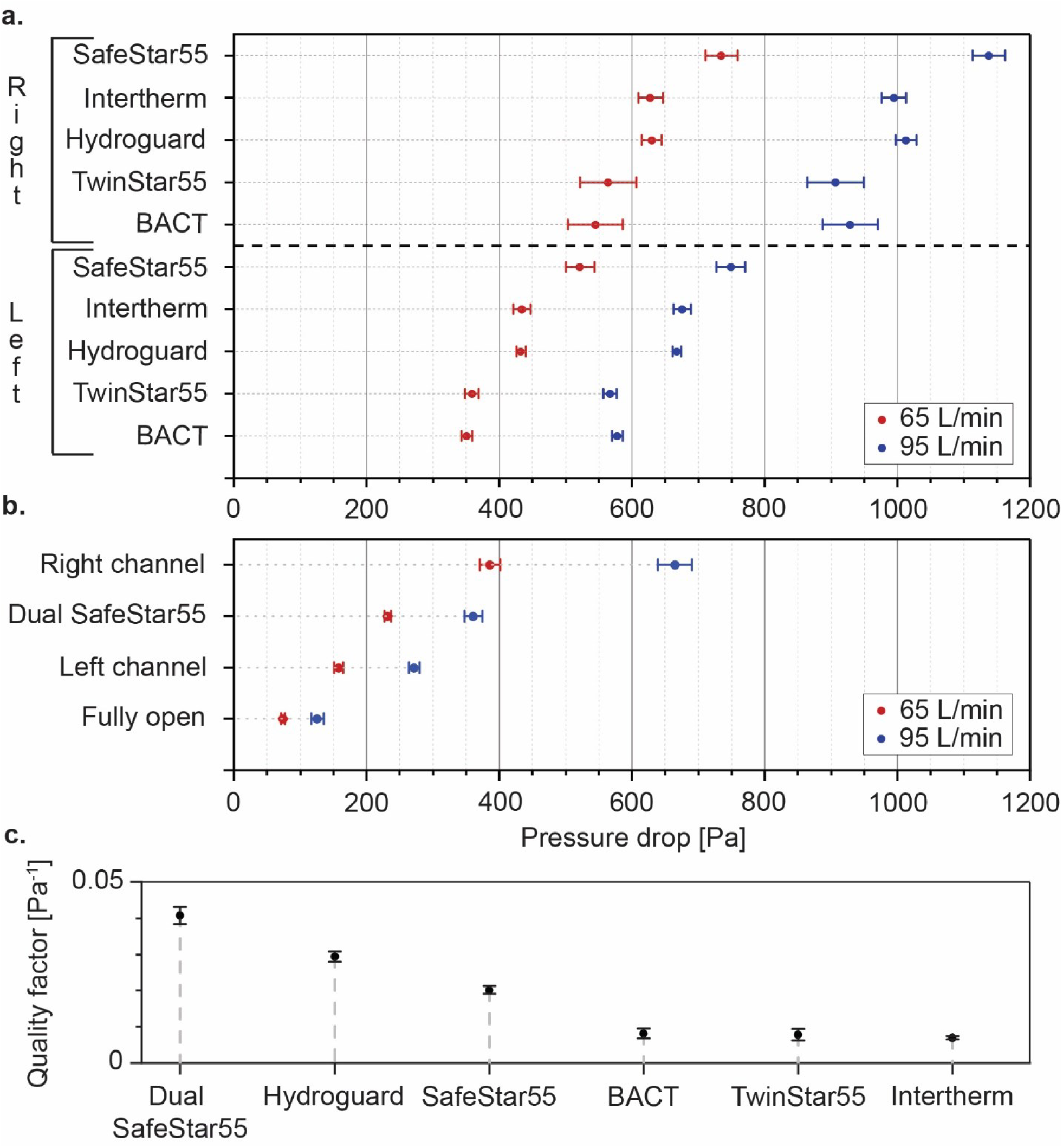
Measurement of the pressure drop and calculation of the quality factor. (a) shows the pressure drop generated by the five medical filters when they are mounted on the face mask with the adapter. The distinction is made between the right and left channel of the adapter (right and left refer to the view shown in figure 1f-g) to highlight the asymmetry of the flow resistance (the other channel being blocked during the measurement). The measurements are performed at 95 L/min to simulate a heavy breathing. 65 L/min corresponds to the highest flow measured through one filter in the dual-filter configuration (a filter connected to each channel) with a 95 L/min pumping flow. It takes into account the asymmetry of the flow resistance between both channels (b) gives the data for the adapters and the dual-filter configuration, as used for the subsequent measurements. Each measurement is repeated five times and the error bars correspond to the standard deviation. In (c) the filters are ranked according to their calculated quality factor based on the filtration at MPPS and their pressure drop at 95 L/min (after subtracting the pressure drop in each channel). The error bars are calculated by combining the error in the filtration efficiency and in the pressure drop measurements.

The pressure drop is lower for a flow of 65 L/min compared to 95 L/min for all filters and configurations, and the left inlet generates a lower pressure drop than the right one. This difference can be explained by the geometry of the full-face mask and the adapter: the left inlet is directly connected to the upper chamber and the flow resistance is lower compared to that of the side channels of the mask, connecting the lower chamber to the right inlet. The use of the SafeStar55 in a dual-inlet configuration (a filter connected to each port) significantly reduces the pressure drop compared to the single-inlet configuration (a filter alternatively connected to the right and left port, the other one being sealed). The Hydroguard and SafeStar55 have significantly higher quality factors than the other filters. As expected, the dual-inlet configuration based on the SafeStar55 shows the highest quality factor as this configuration benefits form a significantly reduced pressure drop. Having a lower pressure drop than the SafeStar55, the Hydroguard would display a higher quality factor in a dual-inlet configuration but due to filter availability, only the SafeStar55 is considered for the further measurements on the full-face mask. SEM images from the filtering material of the SafeStar55 filter are presented in Figure 4. The filter has multiple layers of fibers and a large variability in the diameters of the fibers, measured between 120 nm and 8 µm. Further images are presented in Figure S3.

**Figure 4.**
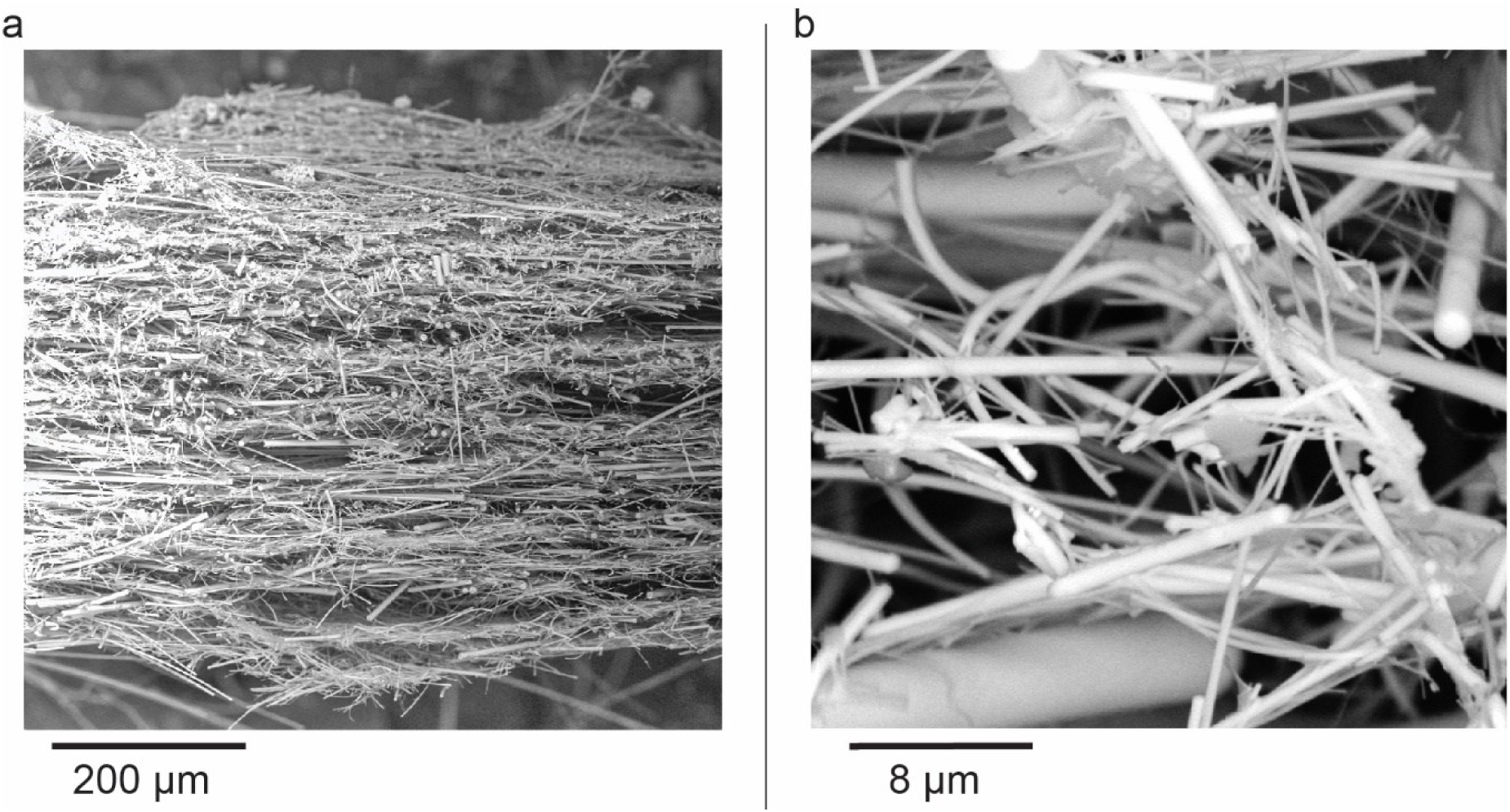
SEM images of the cross-section of a piece of filtering material from the SafeStar55 filter, showing the multiple layers of fibers (a) and the variability in the diameters of the fibers (b).

### Size-dependent particle penetration in the full-face mask

The assembly composed of the 3D-printed adapter and the filters is checked for potential leakages before measuring the particle penetration in the complete mask. Due to the limited spatial resolution of the 3D-printing process, the adapter has rough surfaces and micro-sized gaps can form at the interfaces with the filters in the absence of a sealing gasket. However, the measurements show a particle penetration of 0.2%, which is acceptable. More details are given in Figure S4. On the other end of the adapter, at the interface with the mask, the tightness is realized by a sealing gasket located on the port of the mask.

The SafeStar55 filter is measured at 95 L/min (corresponding to a face velocity of 67 cm/s) to be directly compared with the full-face mask. The setup for the characterization of the filter and the full-face mask in the dual-filter configuration is shown in Figure 5a. The results are presented in Figure 5b. The increased airflow leads to a decrease of the MPPS of the SafeStar55 compared to the data at 8.5 cm/s, from 250 nm to around 50–80 nm. A decrease of the MPPS with increasing face velocity has been previously observed [16, 17] and theoretically explained [18]. The higher face velocity decreases the efficiency of the diffusion-driven filtration, resulting in a higher penetration for particles below 100 nm. Simultaneously, the higher flow increases the efficiency of the inertial impaction leading to a decrease of the penetration of particles above 100 nm. More details on the influence of the face velocity on the filter are given in Figure S5.

**Figure 5.**
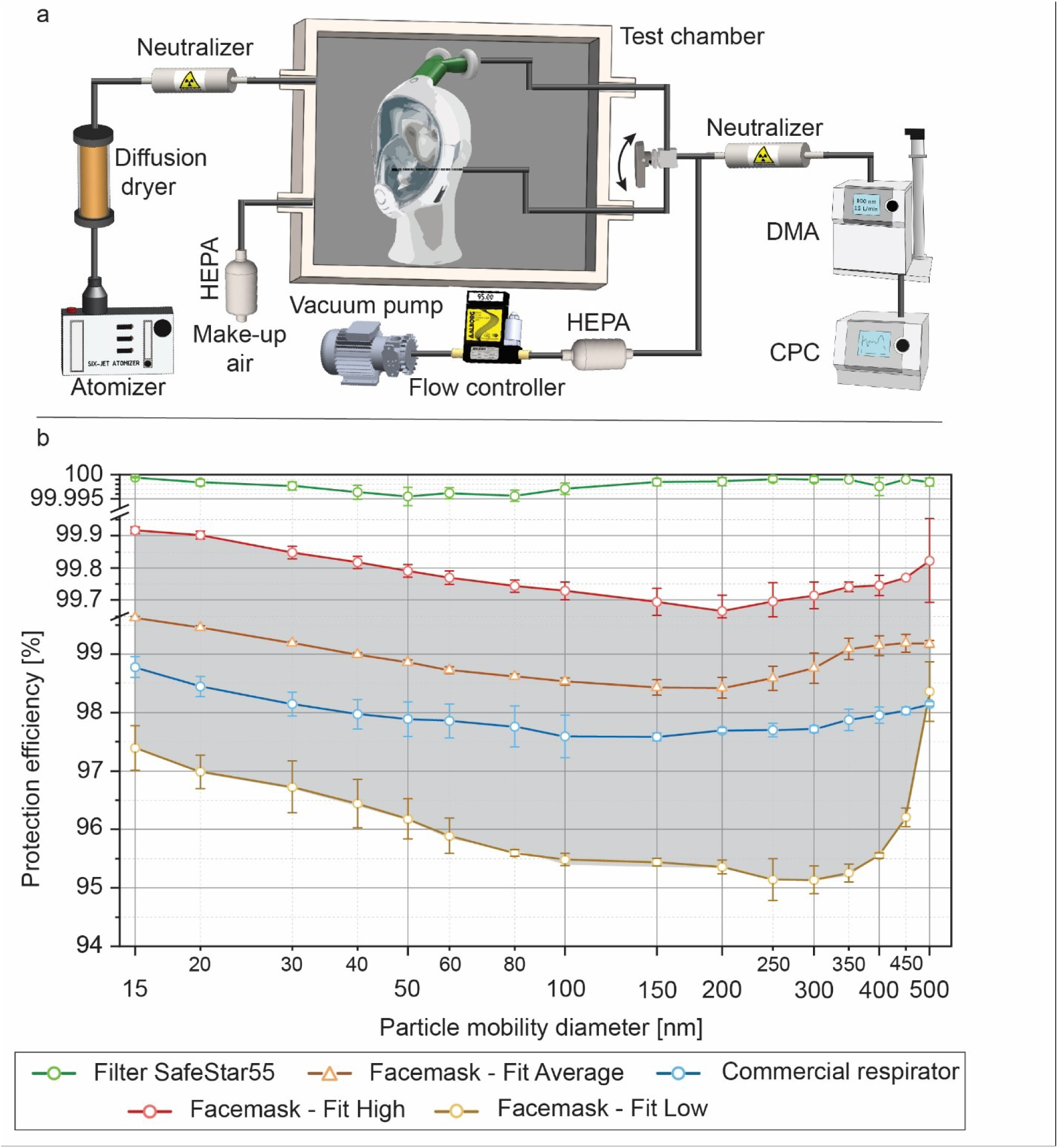
Measurement of the protection efficiency (calculated in the same way as the filtration efficiency and used to characterize the complete mask) of the mask in different fitting configurations(a) shows the experimental setup, with the full-face mask mounted on a polystyrene dummy head and installed in a stainless steel chamber. (b) presents the results of the protection efficiency against the penetration of NaCl particles with different mobility diameters at 95 L/min and different fittings on the dummy head to represent the extreme cases. The grey area represents the range of filtration efficiency depending on the fitting scenario. It is compared to a commercial full-face respirator (3M Model 6800 equipped with two filters A2B2E2K2P3 R, corresponding to a P3 particle filter and stopping 99.95% of the particles according to the EN 143 standard). The filtration efficiency of the SafeStar55 filter is measured at 95 L/min. The error bars relate to the standard deviation calculated from three repetitions of 60 measurement points.

The characterization of the full-face mask is done in the dual-inlet configuration with two SafeStar55 filters. The expression protection efficiency is used for the full-face mask to make the distinction with the filtration efficiency characterizing the filters. They are both calculated with the same method.

In order to take into account the variability due to the fitting on the polystyrene head, three different configurations representing the extreme cases are tested:

- the upper curve marked “High” is the maximal measured filtration efficiency and is obtained after sealing the interface between the polystyrene head and the full-face mask with silicone sealant. This case represents the highest achievable protection efficiency of the face mask and would correspond to a perfect fit of the mask on the wearer’s head.
- the lower curve marked “Low” is measured after the seal check (details on the seal check are given in the section Materials and Methods) is intentionally not fulfilled: the full-face mask is installed on the dummy head but the seal check still shows the presence of leakages which are deliberately not corrected. The purpose of this scenario is to estimate the lower boundary of the protection efficiency, considering that a fulfilled seal check will lead to a higher protection than the configuration tested in the present case. Even if the exact level of leakage is difficult to reproduce, the protection efficiency is higher with a fulfilled seal check than in this scenario.
- the middle curve marked “Average” is representative of the most frequently measured efficiency. This corresponds to a typical fit on the wearer.

The resulting grey area indicates the protection efficiency range depending on the quality of the fitting. It encloses an area between 95% and 99.9% efficiency. The MPPS is around 200 nm for the best and average cases, increases to 300 nm for the lower boundary. The efficiency does not go below 98% when the seal check is meticulously followed. The protection performances of the full-face mask are close to those of the commercial respirator taken as a reference. The measurements highlight the good performances of the full-face mask and the large variability induced by the fitting, and therefore the importance of performing a fitting test before using the mask.

To compare the results obtained on a static polystyrene head with the protection efficiency in realistic operational conditions, the mask with filters (2xSafeStar55) and adapters is tested according to the EN 149 standard requiring movements of the head during the measurements. Three fitting tests are performed with one mask and three volunteers, resulting in particle penetrations of 2.6%, 4.8% and 5.6% with paraffin particles, fulfilling the requirements for FFP2 masks. The measured values are slightly lower than the values presented in Figure 5b. This difference can be explained by the test method, as EN 149 requires the wearer to move during the measurements. These movements create small gaps between the skin and the mask and increase the particle penetration. A comparison of the particle penetration in the mask is done with the dummy head, a non-moving volunteer and a volunteer reproducing the movements of the head as described by EN 149. The results are given in Figure S6 and show a significantly higher particle penetration when the volunteer is moving.

### Decontamination process

Measuring the impact of decontamination processes on the protection efficiency is critical to assess the reusability of the protective equipment and influences its potential to be successfully used in medical facilities. Autoclaving and immersion into ethanol (70 vol.% ethanol in deionized water) are selected due to their availability in most medical facilities. The full-face masks are decontaminated without the adapter and filters, since these components are easily replaceable at a reduced cost. The results are presented in Figure 6.

**Figure 6.**
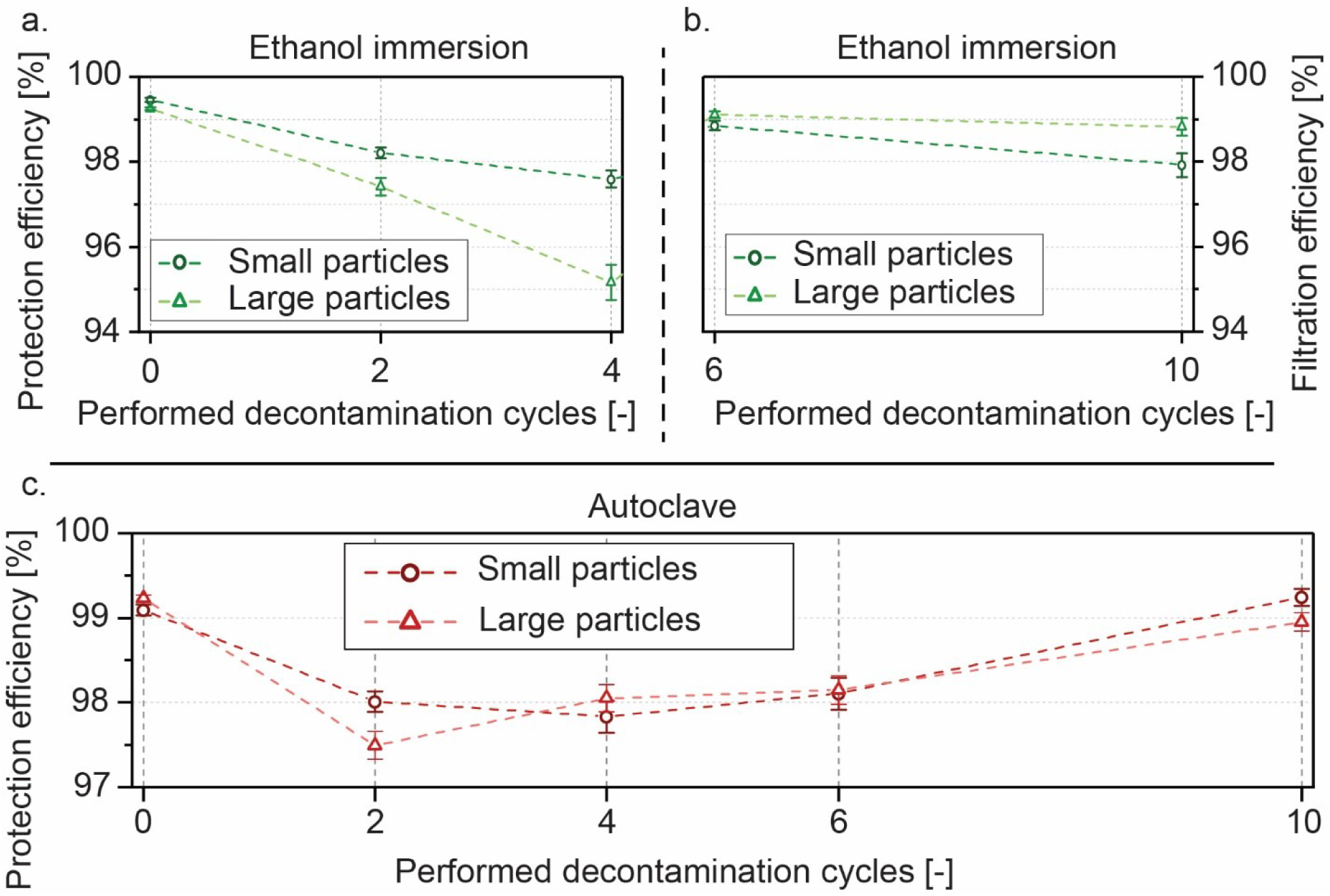
Evolution of the protection efficiency after multiple cycles of decontamination. The sample tested in (a) and (b) has been immersed in ethanol. The drying time before the measurement is 2 h at room temperature for (a) and is increased to 12 h in (b). (c) displays the protection efficiency of the autoclaved mask. Small particles correspond to a polydisperse aerosol centered at 36 nm, and large particles to a polydisperse aerosol centered at 82 nm.

The immersion into ethanol initially causes a drop of protection efficiency (Figure 6a), more significant for larger particles (82 nm peak size) than for smaller particles (36 nm peak size). A visual inspection does not reveal any deformations or damages on the mask. Some polymers are subject to swelling through uptake of ethanol after several days of immersion [19, 20]. The face mask is made of different types of polymers: the transparent front cover of the mask is made from acrylonitrile butadiene styrene (ABS) and polycarbonate, the frame is of polypropylene and the sealing skirt is of silicone. According to [21], a solution of 70% ethanol has little effect on the mechanical properties of silicone. Ethanol does not cause a significant swelling of polypropylene films [22], polycarbonate [23] and ABS [24]. Little information is available on the absorption rates of ethanol, though [25] gives the rates for water absorption by ABS, polypropylene and polycarbonate (< 0.001% dimensional change after 24h exposure of polycarbonate). Another factor can cause the drop of protection efficiency: it is observed during the measurements that the straps used to fit the mask on the head expand more when they are still soaked with ethanol after 2 h drying. This can lead to degraded fitting and therefore to a higher particle penetration. Consequently, the drying time is increased to 12 h (Figure 6b) for cycles Nr.6 to Nr.10 and the efficiency recovers close to the starting value. The straps are completely dry 12 h after being removed from the ethanol solution.

The autoclaved mask (Figure 6c) does not show a significant degradation of its protection performances. The variations can be explained by different drying times between the end of the autoclaving and the measurement. Several damages are detected on the mask: the piece of polymer separating the upper from the lower chamber deforms after two cycles and has to be removed. A gap between the transparent front cover and the frame is also visible, due to the different thermal expansion coefficients of the materials used in both parts. During the removal of the mask from the autoclave after cycle Nr.6, the two pieces separate and the mask has to be reassembled manually. Pictures are shown in Figure S7. However, it does not affect the protection efficiency after cool-down.

The decontamination methods do not cause any significant and permanent degradation of the protection efficiency. Nonetheless, autoclaving leads to permanent mechanical deformations that can severely impact the particle penetration if the mask is not handled carefully. The masks should be given enough time (>12 h) to properly dry or to cool down.

## CONCLUSION

This study analyzes the potential of a modified snorkel mask to be used as PPE. First, the filtering efficiency of various medical filters is measured in comparison to FFP2 and FFP3 filter materials. The pressure drop of the medical filters is measured and the quality factor is calculated. Based on the results, the filter model SafeStar55 is defined as the optimal filter within the framework of this study and used for further testing of the mask. Second, the size-resolved particle penetration in the complete mask is measured. It shows that the mask performs comparable to commercial respirators and the largest source of variability is the fit on the wearer’s head. Measurements according to the EN 149 standard show a compliance with the FFP2 protection level. Lastly, the robustness of the equipment subjected to decontamination cycles with 70% ethanol and autoclaving is determined. The mask shows little change in protection efficiency over ten cycles, if enough drying or cooling time is allowed between decontamination and measurement.

The full-face masks described in this work are not advised to replace specifically designed respirators but constitute a decent emergency alternative in the event of a shortage of FFP2 protections, face shields and full-face respirators to help protecting medical staff exposed to highly contagious patients. The snorkel masks efficiently cover the face, especially the mouth, the nose and the eyes, and reduce the risk of contamination through airborne particles or larger droplets emitted by patients. They only require minor adjustments and are reusable after decontamination without lowering their ability to stop particles. The required adapters are available as an open-source design and can quickly be 3D-printed on site using commonly accessible filaments. The chin valve can be easily and effectively sealed using materials available in most hardware stores.

The mask can be further adapted to better meet the requirements for use in medical facilities: a thinner transparent front cover could reduce the weight and provide better optical properties in order to increase the wearer’s comfort.

## MATERIALS AND METHODS

### Filter selection

The setup for the evaluation of the particle penetration in the filters is described and used in other research [12, 26, 27]. Particles are generated from a solution of NaCl in deionized water by a single-jet atomizer (Model 3079A, TSI Inc., USA). A 0.01 wt.% NaCl solution is used to generate particles smaller than 100 nm and a 1 wt.% solution to generate particles larger than and including 100 nm. The penetration is measured for particle sizes between 12 nm and 300 nm. Although the smaller particle sizes are not relevant to evaluate the penetration of the SARS-CoV-2 (viral particles between 60 nm and 140 nm [6]), they are used here for two reasons: other viruses have smaller sizes (Picornaviruses between 22 and 30 nm, [28] and Parvoviruses between 18 and 26 nm [29]) and the characterization of the filters requires the determination of the MPPS.

The polydisperse aerosol flow is set to 1 L/min before entering the differential mobility analyzer (DMA, Model 3081, TSI Inc., USA) working with a sheath flow of 10 L/min. The resulting monodisperse aerosol exits the DMA and the final flowrate is reached by mixing the aerosol with make-up air. Two condensation particles counters (CPC Model 3775, TSI Inc., USA) sample the air upstream and downstream of the filter. The measurements of the FFP2 and FFP3 filters are performed with a piece of the filtering material having a diameter of 50 mm mounted on a filter holder, the medical filters are measured as received (diameter of the filtering material: 55 mm) and are directly connected to the tubing. The airflow downstream of the filter is treated by a HEPA filter and evacuated by the vacuum pump. The air stream is controlled by a mass flow controller (GFC37, Aalborg Instruments & Controls Inc., USA) in order to maintain a constant face velocity of 8.5 cm/s through the filtering material. It is decided to use a constant face velocity for all the filters in order to allow a direct comparison of their filtration efficiency. For the subsequent testing of the complete full-face mask, the face velocity is adapted to the chosen filter in order to reflect the operating conditions. The sampling time is 5 min, with one measurement every second.

The filtration efficiency is calculated according to the following formula, adapted from [30]:

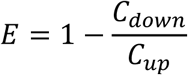

where *C*_*down*_ and *C*_*up*_ represent the particle number concentration up- and downstream from the filter.

The pressure drop is measured on the complete mask, with adapter and filters, installed on a polystyrene head. The setup is described in Figure S8 (pressure sensor PX 409 Series, Omega Engineering Inc., USA). The high pressure point is taken outside of the mask, between the two filters. The low pressure point is measured inside the lower chamber of the mask. The airflow through the mask is generated by two vacuum pumps and controlled by four flow controllers in order to reduce the pressure drop in the system. 65 L/min and 95 L/min flows are generated to represent the most challenging conditions met by the filters in a dual-filter configuration. As only one specimen is available for most of the filters, each one is alternatively installed on the left and the right inlet, the other one being sealed.

A seal check, adapted from [31] is performed before all the measurements:

- The airflow is set to 95 L/min
- The inlet of both filters is obstructed
- A significant movement of the mask towards the head should be observed, indicating an aspiration of the mask caused by the pressure drop and therefore the absence of leakage
- If no movement is observed, the interface between the mask and the head is examined for potential leakages and the mask readjusted until the previous criteria is fulfilled.

Each measurement is repeated five times.

The quality factor is calculated using the following equation [32, 33, 34]:

*Q* = −ln(*P*)/∆*p*

P is the particle penetration and ∆p the pressure drop. The calculation is done by using the penetration at the MPPS and the pressure drop at 95 L/min after subtracting the pressure drop generated by the mask without filters.

The tested filters are obtained from medical facilities and details are given in Table 1. Pictures of the filters are presented in Figure S9.

**Table 1:** Information on the filters tested

| Manufacturer | Name | Filter type | Intended use |
| --- | --- | --- | --- |
| UVEX | Silv-air C | FFP3 | Face mask |
| HD Medis | - | KF94 | Face mask |
| Dräger | TwinStar55 | HME | Mechanical ventilation |
| Dräger | SafeStar55 | HEPA | Mechanical ventilation |
| PharmaSystems | BACT HME Port | HME | Mechanical ventilation |
| Intersurgical | Hydroguard mini | HEPA | Mechanical ventilation |
| Intersurgical | Intertherm HMEF | HME | Mechanical ventilation |

### Fabrication of the adapters

The design of the adapter derives from the design of Filip Kober (Printable ventilator-free respiratory: Subea Easybreath Mask Adapter, GrabCAD). Specifically, the single filter design is modified to incorporate two viral filters which filter both of the flow channels of the mask.

The part is printed in a vertical orientation (filter inlets positioned upward), using an Ultimaker 3 Extended using PLA, 0.4 mm nozzle diameter, 0.15 mm layer height, 20% infill and a skirt build plate adhesion (20 lines, 0 mm distance). PVA is used as support material, to support overhangs of greater than 45 degrees.

### Influence of the chin valve

The model of snorkel mask used for all the measurements is an Easybreath (Decathlon). The chin valve is sealed with a laser cut 3 mm thick Poly(methyl methacrylate), PMMA, disk fused to the mask with acetone after the removal of the silicone valve. This sealing method follows several requirements to make it compatible with the intended use of the full-face mask:

1. Cheap, simple and fast in order to be applied to large quantities of masks
2. Non-toxic as it will be used inside a closed volume
3. Robust, because it must withstand the various disinfection processes applied to the mask Further details on the other considered sealing methods are given in supporting information (Figure S2).

The particle penetration is measured in two steps: in the first step, the mask is fitted on the polystyrene head, in the second step the mask is worn by a volunteer (whose informed consent is obtained) to follow a real breathing cycle. In both cases the challenging aerosol is taken from the environment and two CPCs sample the upstream and downstream aerosol. The measurements with the polystyrene head are carried out at 65 L/min and 95 L/min. The volunteer targets a breathing frequency of 16 movements per minute. For the normal breathing, the functional residual capacity of the lungs is considered, while the inspiratory vital capacity is considered for the heavy breathing. Both volumes are calculated from [35] and lead to a flowrate of 52 L/min for the normal breathing and 83 L/min for the heavy breathing.

The temperature and humidity values are acquired by four sensors (Model SHTC3, Sensirion AG, Switzerland). Sensors Nr.1 and Nr.2 are installed at each side of the upper chamber, Nr.3 in the lower chamber and Nr.4 outside. The chin valve is tested both in the original configuration and completely sealed as described earlier. The mask is fitted on the volunteer and the seal check previously described is performed before each measurement.

### Potential leakage of the adapter assembly

The filters are assembled on the adapter and integrated in the setup previously described (Figure 2a), the filter holder being replaced with the adapter. Minor adaptations are made to the setup: the penetration is measured for a polydisperse aerosol, without considering the different particle sizes and the challenging aerosol is taken from the environment rather than generated by the atomizer. The particle concentration and size distribution in the room where the measurement takes place can be considered as stable over the sampling time (5 min to complete an upstream and downstream measurements). Two filters SafeStar55 are mounted on the 3D-printed adapter. The upstream and downstream airflows are sampled by two CPCs at 1.5 L/min. The filtration efficiency is measured at both 65 L/min and 95 L/min. Potential methods to improve the tightness are tested: winding of Teflon tape around the filter’s inlet and sealing of the assembly by wrapping it in a piece of rubber.

### Size-dependent penetration in the full-face mask

The size-dependent particle penetration in the full-face mask is measured between 15 nm and 500 nm. The minimum particle size is increased from 12 nm in the filter comparison to 15 nm because the dilution of the aerosol, resulting from the higher face velocity, drastically decreases the upstream concentration for 12 nm particles. The polydisperse aerosol stream is generated by a six-jet atomizer (Model 9306, TSI Inc., USA) at an aerosol flowrate of 10 L/min from a solution of NaCl in deionized water, using the same concentrations as previously mentioned. The aerosol flow is directly mixed with 20 L/min filtered air in the atomizer. The flow passes through a diffusion dryer, a Kr-85 neutralizer and is injected into a stainless steel chamber. At the entrance of the chamber, the aerosol is mixed again with filtered air. The airflow pumped out of the mask is set to 95 L/min, equivalent to a heavy breathing. The sum of incoming airstreams (aerosol and make-up air) is set to be slightly higher than 95 L/min in order to prevent external particles from entering the steel chamber and keep a low background concentration. The background concentration in the chamber is checked at the beginning of each measurement and is lower than 1 particle/cm^3^. The full-face mask is installed on a polystyrene head and the particle concentration is measured from the lower chamber using a sampling line running through the dummy head. The aerosol size distribution is measured by a scanning mobility particle sizer (SMPS, Model 3936L75, TSI Inc., USA). The sampling time is set to 60 s for each particle size and the SMPS is controlled manually. The airflow of 95 L/min is applied during the sampling of both upstream and downstream particle concentrations in order to generate the same pressure drop in both sampling lines and compensate particle losses due to the high flow. The CPC operates at critical flow conditions and compensates the variations of the inlet pressure caused by the pressure drop in the face mask. The diameter and length of the tubing is adapted to minimize the pressure drop and keep the inlet pressure of the CPC in the range recommended by the manufacturer. The influence of the pressure drop through the full-face mask on the inlet flow is therefore negligible.

The commercial respirator used for comparison is a 3M Model 6800 equipped with two filters A2B2E2K2P3 R (P3 particle protection class according to EN 143).

The measurements according to the EN 149 standard are performed in an atmosphere charged with Paraffin aerosol. The aerosol concentration is measured with a Portacount Pro+ Respirator Fit Tester (Model 8038, TSI Inc., USA) outside and inside the full-face mask while the volunteers perform a series of tasks (movement) described by EN 149:

Whilst still walking the subject performs the following exercises:

1. walking for 2 min without head movement or talking;
2. turning head from side to side (approx. 15 times), as if inspecting the walls of a tunnel
3. moving the head up and down (approx. 15 times), as if inspecting the roof and floor
4. reciting the alphabet or an agreed text out loud as if communicating with a colleague
5. walking for 2 min without head movement or talking.

### Decontamination process

The decontamination cycles are performed as following:

- Autoclaving: 22 min at 121°C (1.2 bar) followed by 30 min drying.
- Ethanol immersion: 15 h immersion in a solution of 70 vol.% ethanol in deionized water.

Both methods prove to be efficient to inactivate coronaviruses: [36] proves that a solution of 70% ethanol reduces the infectivity of two types of coronaviruses by 99.9% after 1 min. Autoclaving is a widely used method to sterilize medical devices and a lower temperature of 70°C is already sufficient to inactivate SARS-CoV-2 within 5 min [37].

Each mask is exposed to ten cycles of one decontamination method. In order to have a stable aerosol concentration and size distribution throughout the entire duration of the measurements (several days), the challenging polydisperse aerosol is generated by a six-jet atomizer from a solution of NaCl in water concentrated at 0.01 wt.% and 1 wt.%. The size distributions of the generated particles are centered at 36 nm (0.01 wt.%) and 82 nm (1 wt.%). The size distribution of the challenging aerosol is measured with an SMPS. The data is given in Figure S10. The downstream and upstream concentrations are measured by a CPC while the sample is placed on the dummy head in the stainless steel chamber. The adapter and the filters are removed from the mask during the decontamination cycles. [38] shows that the exposure of filtration material to temperatures < 100°C do not impact their filtration efficiency and might therefore constitute an efficient way to inactivate SARS-CoV-2 on the filters. However, according to [39] using ethanol immersion to disinfect electrostatic filters might degrade their filtration performances through the loss of electrostatic charges.

## ASSOCIATED CONTENT

### Supporting Information

The following file is available free of charge.

Influence of the chin valve on the particle penetration, Tested methods to seal the chin valve, SEM images of the SafeStar55 filtering material, Evaluation of the adapter assembly, Evolution of the filtration efficiency of the SafeStar55 with increasing flowrate, Dispersion of the filtration efficiency for the volunteer and the dummy head, Pictures of the damaged mask after autoclaving, Setup for the measurement of the pressure drop, Photo of the filters, Size distribution of the particles generated by the 1 wt.% and 0.01 wt.% NaCl solutions.(Supporting_Information_Alternative_PPE_MedRxiv.PDF)

## AUTHOR INFORMATION

### Author Contributions

J.S. and J.W. conceived the testing plan, L.J. lead the modification of the face mask, designed and fabricated the adapters and provided the masks and the filters, E.A. defined the sealing method for the chin valve, J.S. conducted the measurements in J.W.’s group as well as the data analysis, J.S. and E.A. contributed to the comparison of the filters. C.G. performed the measurements according to the EN 149 standard, B.M. tested the mask in medical facilities and gave his feedback. All authors have contributed to the manuscript and have given approval to the final version of the manuscript.

### Funding Sources

The authors declare no competing financial interest.

## Data Availability

All data is available in the manuscript and upon request from the authors.

## ACKNOWLEDGMENT

The authors would like to thank the initiative helpfulETH that brought the team together and Darko Kalinić for his contribution to the testing the face mask. The work was partially supported by Innosuisse project 46668.1 IP-ENG “ReMask: Strategies for innovations for Swiss masks needed in pandemic situations”.

## ABBREVIATIONS

PPE: personal protective equipment
CPC: condensation particle counter
SMPS: scanning mobility particle sizer
MPPS: most penetrating particle size
DMA: differential mobility analyzer
HEPA: high-efficiency particulate air
ABS: acrylonitrile butadiene styrene
PMMA: Poly(methyl methacrylate)
HME: heat and moisture exchanger
SEM: scanning electron microscope

## Notes

### Competing Interest Statement

The authors have declared no competing interest.

### Author Declarations

No IRB necessary, informed consent has been obtained from the volunteers wearing the mask.

